# Improving Clarity and Interpretability of Items in a Bilingual Index of Propensity to Integrate Research Evidence into Clinical Decision-Making in Rehabilitation

**DOI:** 10.1101/2024.08.07.24310514

**Authors:** Jacqueline Roberge-Dao, Nancy Mayo, Annie Rochette, Keiko Shikako, Aliki Thomas

## Abstract

**Purpose:** To contribute evidence for the clarity and interpretability of a new five-item bilingual multidimensional index of a rehabilitation clinician’s propensity to integrate research evidence into clinical decision-making.

**Methods:** This study was conducted in three sequential steps: (1) We conducted a focus group with occupational therapists, physical therapists, and researchers to review the items and response options for clarity, consistency, and interval properties and agree on equivalency in English and French. (2) We conducted cognitive interviews whereby clinicians elaborated on their interpretation of the item, comprehensibility of items, and appropriateness of response options. Accepted modifications were integrated and tested with subsequent participants. (3) We conducted an online survey to validate the English and French equivalency of response options on a 0 to 100 scale.

**Results:** During the qualitative revision process (one focus group with 7 participants followed by 27 interviews), the index was revised 12 times with substantial modifications to the *use of research evidence* and *attitudes* items.

**Conclusion:** This research increases the clinical relevance and reduces measurement error of this brief index which can inform on individual or organizational factors influencing a clinician’s propensity of integrating research evidence into decision-making and ultimately, improve rehabilitation outcomes.

## Introduction

To optimize quality of care, occupational (OTs) and physical therapists (PTs) as rehabilitation clinicians are expected to engage in evidence-based practice (EBP), that is, integrate best available research evidence, their clinical expertise, and patient values and preferences when making clinical decisions.^1–4^ Rehabilitation clinicians acknowledge that clinical experience is essential to integrating research evidence into practice^13–15^ and as such, they must make sense of the quality, pertinence and applicability of research evidence using their judgment and tacit knowledge.^16,17^ Importantly, patient-centered practice is considered a basic tenet of occupational therapy (OT) and physical therapy (PT) and accommodating patients’ goals, values and preferences are vital for positive patient outcomes and satisfaction.^18,19^

The tripartite conceptualization of EBP (i.e. research evidence, clinical expertise, and patient values) has traditionally been depicted by the “three circles” model.^5–8^ In more recent years, there have been refinements and additions to this conceptualization of EBP reflected in the inclusion of the context (e.g., organizational, economic, professional) to highlight the external influences to clinical decision-making (CDM).^9–12^ Despite the purported benefits of such a CDM approach and the implementation of EBP content into most entry level OT and PT curricula^1,4^ globally, clinicians report difficulties integrating research evidence into practice.^20–25^ Lack of allotted time for activities related to EBP, poor access to research evidence; low confidence in applying research to practice; lack of knowledge that evidence-based interventions exist; and inadequate equipment to implement new practices are among the reasons for the underutilization of research evidence in practice.^15,20–22,25–27^

Robust measurement practices are needed to identify the factors related to EBP that should be improved or the strong areas that must be maintained.^28–30^ Identifying which areas require improvement can inform targeted allocation of resources to support EBP and ultimately, improve patient outcomes. There exists a vast selection of questionnaires measuring the core factors influencing an OT or PT’s likelihood to integrate research evidence.^28,31–33^ However, there are shortcomings to current EBP measures such as failure to concurrently measure multiple domains, the inappropriate analysis of items derived from ordinal scales and the unknown relative weight of domains.^33^

In our previous work, Al Zoubi et al. identified the six most salient domains influencing a rehabilitation clinician’s likelihood of integrating research evidence into CDM.^30^ One best performing item was chosen per domain to form a brief, multidimensional index in English and French, as described by Roberge-Dao et al. (submitted). The five domains included in the index are: *use of research evidence, self-efficacy, resources, attitudes,* and *activities related to EBP.* However, as the selected items stem from five different questionnaires, there is inconsistency between items (and response options) in terms of the terminology and formulation which can increase respondent burden and introduce measurement bias. In addition, the English and French versions of these items may present cultural or linguistic discrepancies that can introduce systematic differences in scores.^34^

The aim of this study was to contribute evidence for the clarity and interpretability of items and response options for a new bilingual measure, the *Propensity to Integrate Research Evidence into Clinical Decision-Making Index* (PIRE-CDMI). Specifically, the primary objective was to review and revise the included items in the prototype index in English and French. The secondary objective was to estimate the equivalency of response option labels in both languages.

## Methods

This study involves a three-phased qualitative review process as illustrated in figure 1. Ethics approval was obtained from *The Faculty of Medicine and Health Sciences Institutional Review Board* at McGill University for all phases of this study before commencement and recruitment.

**Figure 1.**
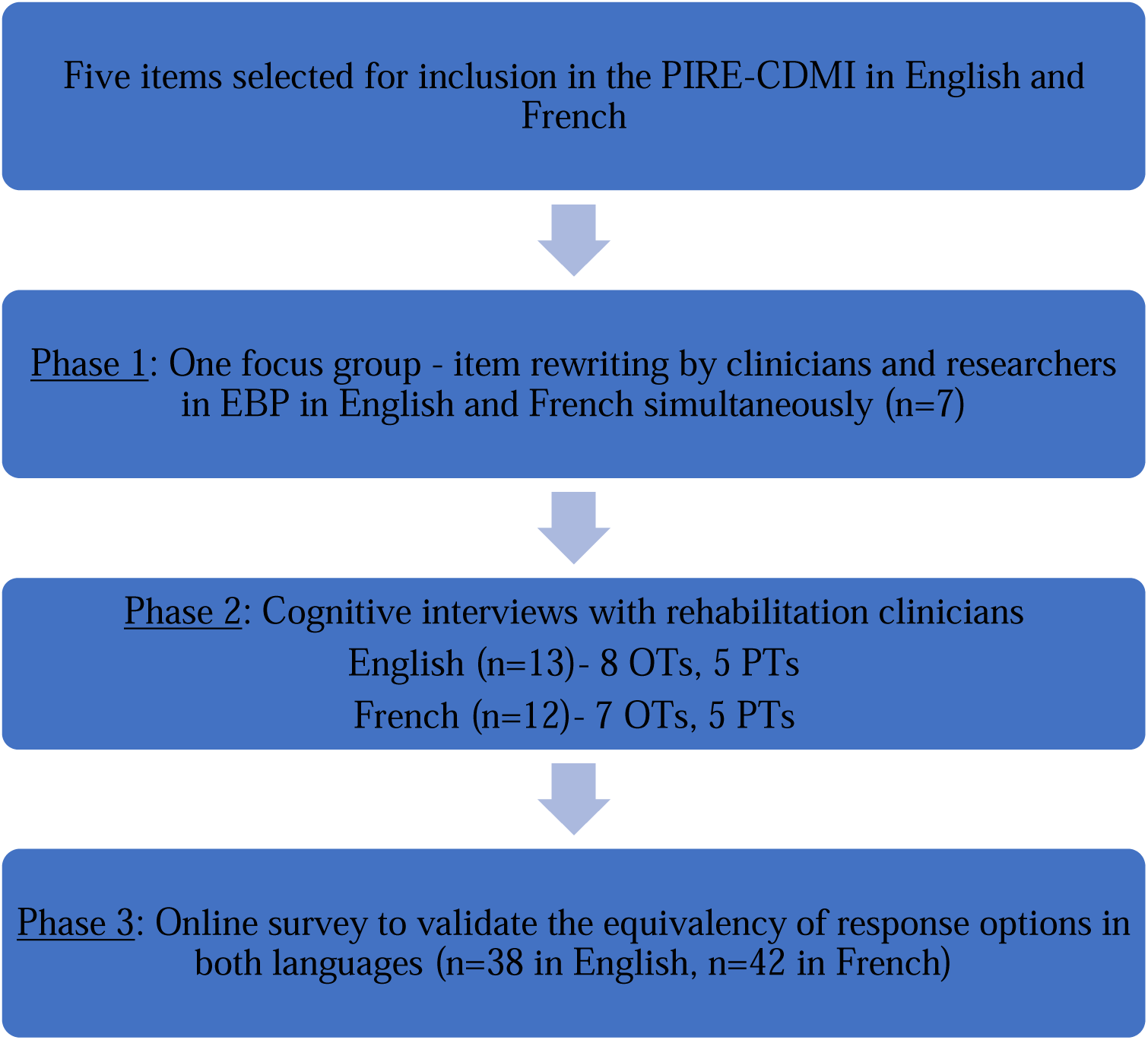
Overview of the item revision process for the Propensity to Integrate Research Evidence into Clinical Decision-Making Index (PIRE-CDMI)

### Phase 1: Focus group

#### Focus group participants

Practicing OTs and PTs and EBP researchers (defined as researchers having experience in EBP research and having published a minimum of one EBP-related publication) were recruited purposefully from the networks of the research team to participate in a 90-minute online focus group. The pool of participants was expected to be bilingual and have equal representation of both French and English native speakers.

#### Focus group process

The aim of this focus group was to review the items and response options of the prototype PIRE-CDMI for clarity, consistency, and interval properties and arrive at a consensus on modifications that would be needed to have equivalent versions in English and French.^35,36^ Participants were asked to establish equivalence in both languages such that the items, instructions, and response options were conceptually (i.e., do people in both groups see the concept in the same way) and semantically (i.e., the meaning attached to words in an item) comparable.^36,37^ Consenting participants were sent the items with a reminder of the study aim a week before the focus group.

The online focus group was conducted via Zoom and structured as follows: (1) welcoming and overview of the study; (2) objectives, instructions, and an example for item rewriting; (3) breakout room with two individuals per room for five minutes to allow attempts at reviewing one item; (4) attend to any questions that arose during breakout room; (5) item rewriting exercise altogether using the share screen function.

During the item rewriting exercise, the moderator (first author JRD) structured the discussion and a note taker recorded the suggested modifications on a shared online document. Participants were asked to rewrite items from question-item format into declarative statements from the perspective of a clinician (see figure 2 for an example of this process). Probing questions included: (1) How would you rewrite this item into a declarative statement? (2) Is the wording clear, and if not, how would you change it? (3) How difficult would it be for OTs and PTs to answer these items? For each item, the French translation was discussed simultaneously. Once every item was discussed, the moderator asked participants to verify that the overall index was coherent in terms of wording and length, that items read well together and that everyone agreed on the final set of items. After the focus group, the research team (consisting of bilingual EBP researchers in rehabilitation) reviewed the suggested final set of items and resolved any withstanding discrepancies.

**Figure 2.**
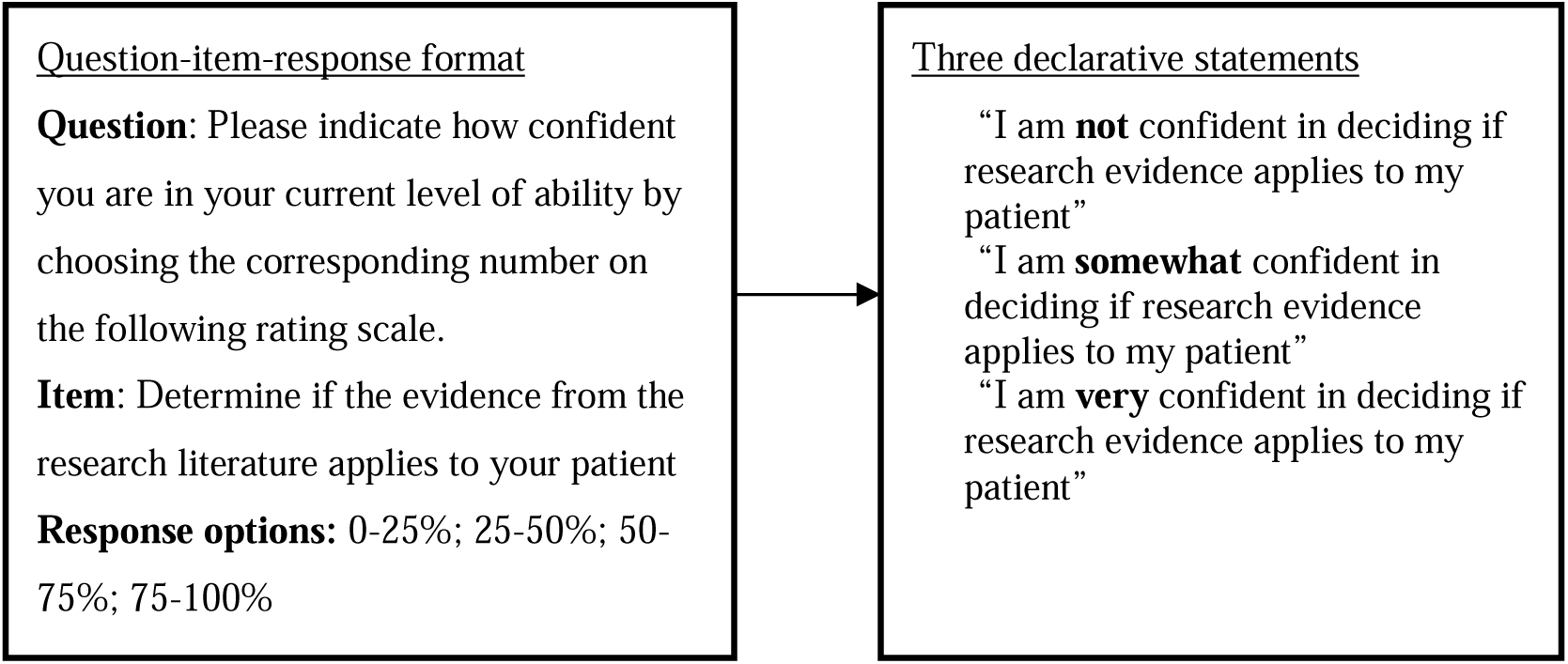
Example of the item rewriting exercise in the focus group

### Phase 2: Cognitive interviews

#### Cognitive interview participants

New participants were recruited for cognitive interviews. Clinicians were eligible to participate in the cognitive interviews if they were (1) practicing OTs and PTs in Canada; (2) native French or English speaking; and (3) had been practicing for a minimum of one year. We used social media (Twitter and Facebook) and the University’s newsletter to advertise the project. Interested participants entered their contact information in an online form. A member of the research team contacted them to provide more information on the study. Participants gave informed consent to participate in the research.

#### Cognitive interview process

Cognitive interviews were conducted by the first author (JRD) by telephone or by Zoom conferencing with potential respondents of the PIRE-CDMI (i.e., OTs and PTs) to identify and rewrite any problematic items to increase the overall readability, functioning and interpretability of the measure.^36,38–42^ All interviews were audio recorded and expected to last between 15 to 30 minutes.

Participants were provided with a copy of the newly reviewed PIRE-CDMI at least one day before the scheduled interview. As presented in table 1, the interviewer used the verbal probing method to elicit participants’ comprehension of all five items by asking specific questions regarding meaning, clarity, and interpretation of items.^40,43^ These questions were adapted from the authors of a study using similar methods in developing a preference-based index for multiple sclerosis.^44^ Participants were encouraged to think out loud while going through the measure, allowing for insight into how a participant perceived and interpreted the items.^43^

**Table 1:**
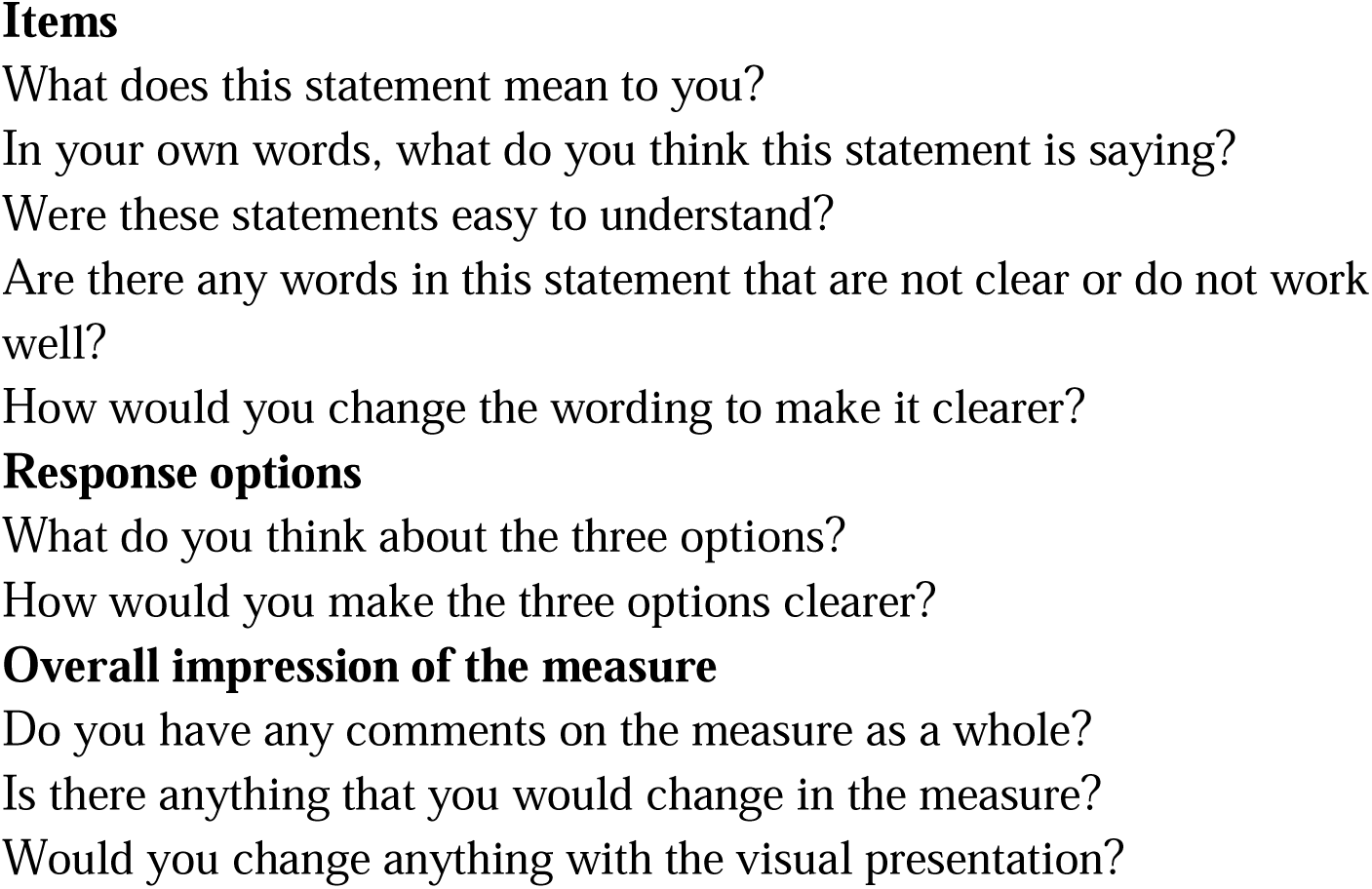
Cognitive interview probing questions

#### Analysis of cognitive interviews

English and French interviews were conducted in parallel so that no language was prioritized. After each day of interviews, the interviewer reviewed comments and revised the problematic items based on participants’ suggestions. The research team reviewed the feedback before implementing the change and proposed suggestions based on best practices of item development such as having simple items that express a single idea, using common vocabulary, and avoiding colloquialisms.^36,41,45^ Changes were implemented in both languages simultaneously, when applicable. The revised version of the PIRE-CDMI was then tested on the next round of participants. interviews were conducted until no further changes were necessary as suggested by three consecutive participants.^44,46^

### Phase 3: Survey

Given Canada’s linguistic diversity, methods to ensure the equivalence of questionnaire versions in the two official languages are warranted to decrease systematic differences between language groups. Validation of translations through quantitative response scaling can contribute to demonstrating that respondents are interpreting items in a similar fashion and consist of asking respondents to denote the position of response options on a visual analogue scale (VAS) (i.e., a line from 0-100) and to compare the ratings between languages. These methods have been used in previous studies.^47,48^

#### Survey respondents

This phase consisted of a cross-sectional online survey to generate additional evidence on the equivalency of PIRE-CDMI response option labels in English and French. The target population was healthcare professionals and students provided that (1) they were native English or French speakers and (2) worked or studied in a healthcare professional or graduate program in Canada. We used convenience sampling and did not exclude respondents based on profession or level of training, because the nature of the survey was such that respondents solely needed to have the abilities to interpret common words and rate response option labels on a numerical scale. We recruited through social media and interested respondents were invited to follow a link with study information, a consent statement, and an invitation to start the survey in the native language of their choice. No identifying nor sociodemographic information data were collected.

#### Survey procedure and analysis

The survey was piloted with seven graduate students, all of whom were also practicing clinicians in rehabilitation. Modifications were integrated to improve survey clarity and task comprehension. The survey was open from November to December 2021.

Each PIRE-CDMI item was associated with three response option labels. Respondents were asked to indicate the position of each of the three response option labels on a 0-100 VAS between two anchors. Response option labels belonging to the same set appeared on a single page sequentially. This method has been previously reported for health-related quality of life measures such as the SF-36^47^ and EuroQol-5d^48^. Specifically, participants were asked: “On the line, where would you position each of the three response option labels between [the bottom anchor] and [the top anchor]?” Appendix I presents the three response option labels for each item and associated response anchors. The full PIRE-CDMI item was also stated on the same page as the response set to provide context.

It was hypothesized that respondents would position the lowest response option label closest to the zero anchor, the middle response option label in the middle, and the highest response option label closest to the 100 anchor. Post hoc analyses were conducted to remove respondents who likely misunderstood the task reflected in having either: (1) more than two sets of disordinal response patterns; (2) the same rating for all three response options for one item or more; or (3) two or more extreme outlier ratings (0 or 100). Multiple linear regression was used to estimate the extent to which VAS ratings per item (0 to 100 scale) depended on language (English, French), response option (low, middle, high) and the interaction between language and response option. The normal probability plot of standardized residuals was visually examined.

## Results

### Focus group item rewriting

Four PTs and three OTs, all doctoral candidates with research experience in EBP participated in the 90-minute focus group. Each of the five question-item-response sets was transformed into five sets of three declarative statements which were then clarified and harmonized in English and French (version 2 of the PIRE-CDMI). Participants agreed that response options including the word “never” (for example, “I never integrate research evidence”) were perceived as being undesirable because choosing these response options would make them be perceived as incompetent. Given that clinicians would not opt for these options, the participants removed the word “never” from the *use of research evidence* and *activities* items. For the *activities* item, focus group participants communicated that it was important not to confine research evidence to scientific articles; they suggested replacing “read research” with “consult research evidence”. Further, participants agreed that omitting the verb “reading” was more inclusive to individuals who may have visual impairments. In French, multiple terms were proposed for “research evidence” but the agreed upon term was “*données probantes”* which was said to be most employed and recognized among clinicians.

Three issues remained unresolved after the focus group and were subsequently discussed within the research team. Modifications were made to the items before starting the cognitive interviews (version 3). First, it was unclear which term was preferred between “patient” and “client” as the terms are often used interchangeably in rehabilitation contexts depending on the setting and the population. The research team agreed to use “patient” consistently and added a footnote to explain the interchangeable nature (N.B. the final wording of items did not include the words “patient” nor “client”). Second, participants could not decide which term was best between “organization” and “clinical setting”. The research team modified the item to focus on the broad availability of resources and refrained from using either term. Finally, participants could not come to a consensus between the verbs “willing” and “inclined” in the *attitudes* item as they were compelled to answer high on the *attitudes* item; although clinicians may be willing or inclined to use evidence, they may not actually do so in practice. Participants reported that the adjectives “willing” and “inclined” did not have equivalent translations in common French that would be suitable for a self-report measure (“*enclin à”* or “*disposé à”* are not commonly used words). Thus, the *attitudes* item was reframed from “I am willing to use EBP” to the notion of “it is worth the effort to [use EBP]”. Appendix II reports the step-by-step changes at each step of the qualitative rewriting process.

### Item modifications from the cognitive interview process

Twenty-four individual cognitive interviews were conducted with 10 PTs and 14 OTs in Canada (13 native English speakers, 12 native French speakers; one bilingual participant provided feedback in both languages). Appendix III presents an overview of the item evolution process during cognitive interviews. An overview of the modifications made to the items are described below.

The *self-efficacy* item underwent three iterations. From the initial item, “I am (very confident/somewhat/not very confident) in my ability to integrate evidence into my intervention plan”, the word “integrate” was replaced with “apply” to be more action oriented. The words “intervention plan” were first replaced with “clinical cases” to avoid discriminating clinicians who solely perform assessments. The words “clinical cases” were then simplified to “practice” to avoid any confusion associated with the variability in clinical cases. Finally, the response option label “very confident” was changed to “confident” because participants stated it was difficult to endorse being very confident with one’s ability to apply research evidence to practice. In the final version, the wording of two response option labels (“somewhat” and “not very” confident) was not an exact translation in French (“*moyennement”* and “*peu confidant”)*.

The item on *use of research evidence* underwent three iterations. At the start of the cognitive interviews, this item consisted of asking respondents about the source of information, between research evidence, colleagues, or clinical experience, that they would seek when faced with a practice uncertainty. Participants found this item particularly difficult to answer because it was dependent on the case at hand (e.g., the availability of evidence for a clinical diagnosis or patient values) and the organizational context (e.g., whether colleagues were available and/or had experience related to the case). Clinicians reported that they often used a combination of all three sources and that it was difficult to select one to describe their typical behavior. The final wording focused on the frequency of using research evidence when faced with a practice uncertainty which avoids the conflicting response options of colleagues and clinical experience. Finally, an asterisk was added to define practice uncertainty as “a situation in which there is a gap in your knowledge relating to a clinical decision”.

With six versions, the *attitudes* item underwent the highest number of iterations. At the start of the cognitive interviews, respondents were asked the extent to which incorporating evidence into practice was worth the effort. Participants suggested that the item not contain the connotation of “worth the effort” because it was (1) prone to social desirability bias (e.g., participants felt pressured to respond the best and highest level) and (2) did not translate well in French (e.g., “*cela vaut l’effort”* or “*cela vaut la peine”*). The item was modified to focus on EBP requiring effort (e.g., “It requires little/some/a lot of effort to integrate research evidence into practice”). The response option label “some (effort)” was changed to “moderate (effort)” to clarify the middle level response, and the words “(requires… effort) for me” were added to clarify the intent of eliciting the individual’s perception of effort rather than a general belief. In French, the direct translation of “it requires little effort for me to…” is “*cela me requiert peu d’efforts pour*…” which was problematic for two reasons. First, starting a sentence in French with “*cela”* was too informal. Second, the verb “*requiert”* was too formal. The structure of the French sentence was changed to place the object (“*intégrer les données probantes dans ma pratique”*) before the verb and to replace “*me requiert”* with “*me demande”*.

The *resources* item underwent three iterations. The initial item was “I feel that I have the/only some of/do not have the necessary resources to integrate research evidence into my practice”. Participants suggested omitting the words “I feel that …” and questioned which resources the item was referring to. An asterisk was added to clarify meaning and enumerate examples of resources facilitating EBP. In French, participants prefered the verb “*je possède (les ressources nécessaires)”* to “*j’ai (les ressources nécessaires)”*. For one response option label (“some of”), the final French wording was not an exact translation (“*une partie des”*).

Finally, the *activities* item underwent the least number of modifications. The only modification consisted of changing the words “consult research evidence” to “keep up to date with research evidence”. Participants were interpreting the initial item as the frequency of using evidence in their practice which was already reflected in the *use of research evidence* item. The revised item reflects the concept of staying up to date with research evidence as an activity outside of routine CDM. Though three of the 24 participants suggested that we explicitly describe and quantify the three adverbs (regularly, occasionally, and rarely), the research team decided to avoid quantifying these adverbs as there is no agreed upon best practice for behavioral frequency of consulting the literature. By providing these three response options without specifying the exact range, the research team intended to capture clinician’s self-report relative to their temporal understanding of keeping up to date with research evidence in their field. In French, “keeping up to date with research evidence” did not exactly translate, so the following modification was retained for conceptual equivalence “*se tenir à jour quant aux données probantes”*.

The initial instructional prompt was “For each group of statements, select ONE statement which best applies to you. Please respond as honestly as possible”. The prompt was modified three times until the final version, “Please select ONE statement from each box which best reflects your current practice and context.”

The visual presentation of the measure was improved following participant suggestions. Specifically, the lettering of each response option was bolded to make discriminating between levels easier. It was also suggested to number the five items (1 to 5) and letter the three response options (a, b, c) to reduce cognitive burden involved in completing the index. The final version of the PIRE-CDMI in English can be found in Appendix IV. The final version of the PIRE-CDMI in French can be obtained from the corresponding author by request.

### Scaling of response option labels

Among the 129 individuals who started the online survey, 60 were Canadian French native (46%) and 69 were English native (54%). Of the 129, 42 Francophones (32%) and 38 Anglophones (30%) were included for analysis. The rest were excluded due to incomplete surveys (n=25, 19%) and task miscomprehension (n=24, 19%). Descriptive results for the rating of the five response option sets by the 80 respondents is presented in table 2. The ordinal nature of the ratings of response sets is illustrated in figure 3. Multiple linear regression results did not suggest any important main effects of language on score for the five items, nor any important interaction of language and response option. Sparring the presence of a few outliers in all items, the residuals were normally distributed.

**Table 2:** Descriptive statistics for the rating of response options on a 0-100 scale (n=80)

|  | Mean (SD) | Min | Max | Mean (SD) in English (n=38) | Mean (SD) in French (n=42) |
| --- | --- | --- | --- | --- | --- |
| **Item 1:** I am _____ in my ability to apply research evidence to practice.<br>**Anchors:** No confidence / Full confidence |  |  |  |  |  |
| Not very confident | 13.8 (5.2) | 0 | 28 | 14.1 (3.7) | 13.6 (6.2) |
| Somewhat confident | 50.6 (6.6) | 30 | 74 | 49.8 (6.3) | 51.3 (7) |
| Confident | 85.9 (5.9) | 70 | 100 | 85.4 (4.7) | 86.3 (6.9) |
| **Item 2:** When faced with a practice uncertainty, I _____ use research evidence.<br>**Anchors:** None of the time / All of the time |  |  |  |  |  |
| Rarely | 11.9 (4.9) | 0 | 26 | 11.9 (3.5) | 11.9 (5.9) |
| Sometimes | 47.7 (7.9) | 25 | 75 | 48.1 (7.1) | 47.4 (8.7) |
| Almost always | 87.3 (5.3) | 70 | 100 | 87.5 (4.1) | 87.2 (6.3) |
| **Item 3:** It requires _____ for me to integrate research evidence into practice.<br>**Anchors:** No effort / Full effort |  |  |  |  |  |
| Little effort | 18.8 (5) | 0 | 27 | 18.6 (4.5) | 19 (5.4) |
| Moderate effort | 52.9 (5.7) | 38 | 70 | 52.3 (3.5) | 53.3 (7.1) |
| A lot of effort | 86.4 (4.8) | 75 | 100 | 87.2 (4.1) | 85.6 (5.2) |
| **Item 4:** I _____ keep up to date with research evidence.<br>**Anchors:** 0 days/month and 30 days/month |  |  |  |  |  |
| Rarely | 4.8 (4.9) | 0 | 17 | 4.6 (4.4) | 4.9 (5.3) |
| Occasionally | 21.3 (11.3) | 3 | 60 | 20.7 (7.8) | 21.7 (13.7) |
| Regularly | 45.9 (18.2) | 13 | 100 | 44.8 (16.2) | 46.8 (20) |
| **Item 5:** I have _____ to integrate research evidence into my practice.<br>**Anchors:** None / All imaginable resources |  |  |  |  |  |
| Few of the necessary resources | 14.1 (4.6) | 3 | 25 | 14.0 (3.8) | 14.2 (5.2) |
| Some of the necessary resources | 43.4 (6) | 20.0 | 56.0 | 43.8 (6.5) | 44 (9.3) |
| The necessary resources | 79.2 (7.2) | 60.0 | 100.0 | 79.5 (7.2) | 79.3 (8) |
| SD, standard deviation |  |  |  |  |  |

**Figure 3.**
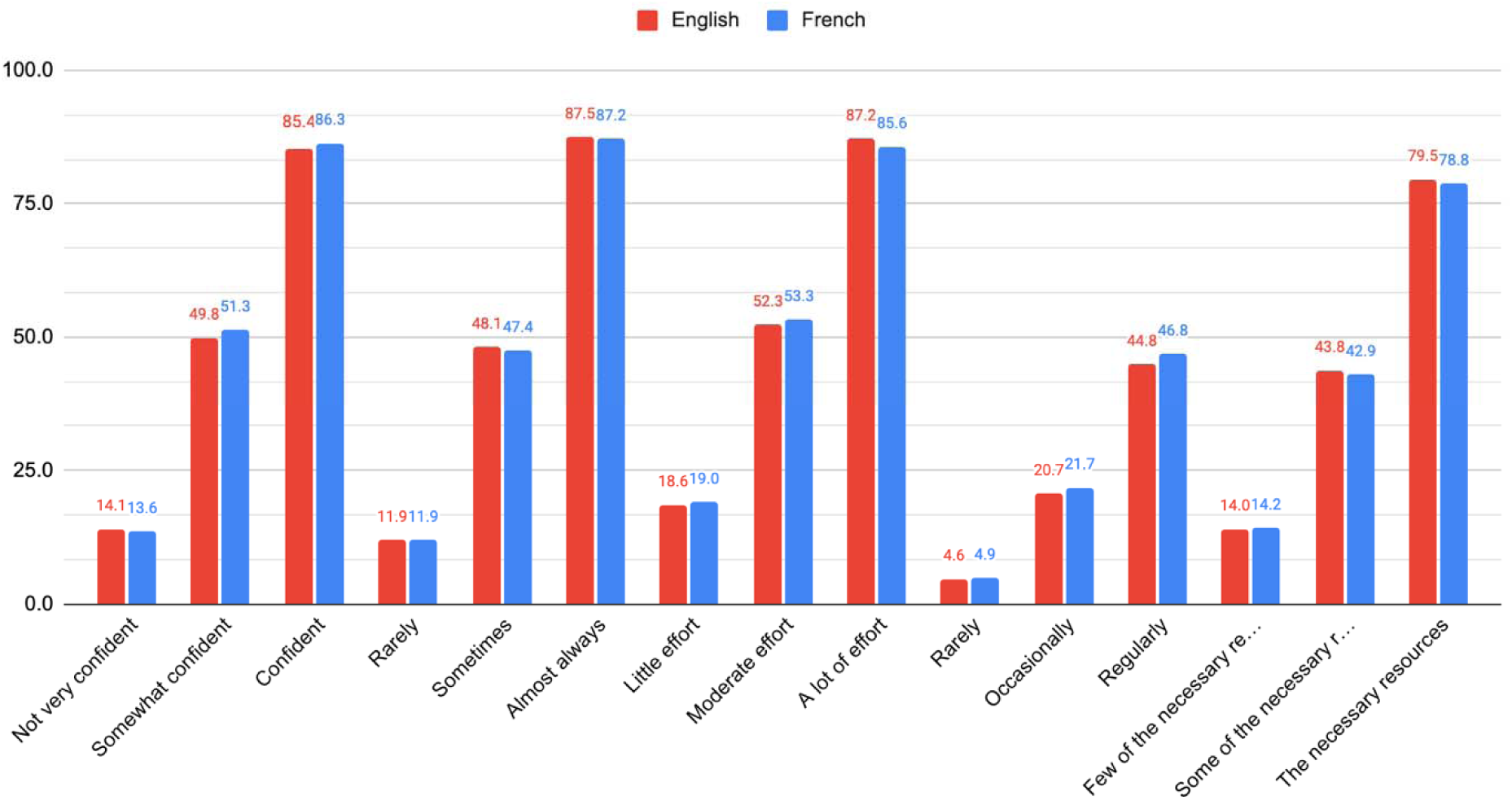
Histogram illustrating the frequency distribution of mean ratings of response option labels on a 0-100 scale for the five PIRE-CDMI items in English and French

## Discussion

This study describes a robust item revision and rewriting process of an index measuring a rehabilitation clinician’s propensity to integrate research evidence into CDM. This three-step approach to qualitative revision has not been previously reported in the EBP literature. Overall, the index (PIRE-CDMI) underwent 12 iterations to increase the clinical relevance and reduce measurement error with important changes made to the *use of research evidence* and *attitudes* items.

The *use of research evidence* item changed considerably with early iterations relating to (1) the frequency of integrating the three EBP pillars (research evidence, clinical expertise, and patient preferences) and (2) clinicians’ primary source of knowledge (research evidence, colleagues, and clinical experience). These two iterations failed to produce useful information because clinicians attested to integrating all three pillars of EBP into CDM and relying on all proposed sources of knowledge to various extents. In fact, the tripartite definition is foundational to how rehabilitation clinicians conceptualize EBP.^50^ However, asking clinicians to select their most relied upon pillar of EBP or to determine the frequency at which they integrate the three components is anathema to the reality of CDM whereby these elements are inextricably intertwined.^23^ Further, asking clinicians to select the most relied-upon source of knowledge in a measure relating to EBP appears to introduce high levels of social desirability bias.^51,52^ For example, a respondent may interpret the desirable answer to be “research evidence” and the undesirable answer to be “colleague” or “clinical experience”. This item formulation could inadvertently imply that consulting a colleague or relying on clinical experience is ill-advised when, in fact, these sources of knowledge are foundational to being a competent, reflexive, and evidence-based clinician.^17,24,50,53^ Though plurality of knowledge in CDM is increasingly invoked in the field of EBP,^54^ rehabilitation clinicians continue to voice challenges related to the integration of research evidence into CDM signaling a deep-rooted need for support with this component of EBP.^21,24,26,27^

The notion of a clinical uncertainty was introduced to the *use of research evidence* item to contextualize the behavior of seeking and using research evidence in CDM. The addition of contextual cues in items has been said to increase the validity of responses, notably when the behaviour has an element of automaticity.^55^ CDM often relies on automatic and intuitive reasoning rather than analytical reasoning, a phenomenon which is hypothesized to become stronger over time.^56,57^ This clinical uncertainty allows a clinician to tap into their analytical reasoning and can be compared to *the event* proposed in the reflective practice literature which is defined as “an event that occurs in everyday practice […] that leaves the occupational therapist with the urge to revisit it to make sense of it for the benefit of his or her future practice”.^58^(p345) The need to nuance this item with a clinical uncertainty is further reinforced by a possible mechanism whereby, over time, research-based knowledge becomes consolidated into tacit or experiential knowledge; in such cases, it may no longer be distinguished as *research evidence* but rather transformed into expert practice that is adapted to the practice context.^59,60^ Correspondingly, it may be difficult for clinicians to discern how frequently they use research evidence on a day to day basis. Lastly, consulting research evidence in everyday practice may not be realistic nor be a desirable behavior as it could conceal other professional difficulties such as low confidence in one’s clinical reasoning abilities.^61^ Thus, the more compelling question is not so much whether clinicians consult formal sources of evidence every day, but whether they do so when confronted with a gap in their knowledge.

Many modifications were made to the *attitudes* item to remedy the social desirability bias reported by participants. Despite changes to the item, participants, most of whom have been educated in Canada, continued to report that EBP was a desirable process and were compelled to select the highest response option for attitudes. The evidence demonstrating the relationship between attitudes towards EBP and EBP behavior is inconclusive. While some studies have suggested that holding positive attitudes towards EBP is an important precursor to EBP behavior,^63–67^ others have demonstrated that they do not translate into effective EBP behavior.^20–22,68–71^ We postulate that measuring attitudes towards EBP may not be useful in the context of this brief index given that (1) the relationship between attitudes and EBP behaviors is uncertain; (2) it is well-established that rehabilitation clinicians are generally convinced of the value of EBP and believe it to be a desirable and necessary process;^20–22,69,71–73^ and (3) value-laden items which can prejudice respondents should be omitted from measures^36^ and attitudes are inherently value-laden. Furthermore, when assessing attitudes for predicting behaviors, it is recommended to avoid measuring attitudes towards a general concept and to focus on specific behaviors.^74^ Using *effort* instead of *attitudes* circumvents asking clinicians whether they consider EBP to be valuable and highlights the perceived cost of integrating research evidence into practice.^75^ *Effort* is defined by the Cambridge Dictionary as “physical or mental activity needed to achieve something”. Social psychology and behavioral theorists have identified effort as largely contributing to behavioral motivation.^76–78^ People are less likely to engage in a behavior if it requires a large amount of effort^79^; several studies have shown that rehabilitation clinicians perceive the enactment of EBP as being effortful.^23,24,69^

One noteworthy modification to the *activities* item is the inclusion of various sources of research evidence outside of scientific articles. As such, one could associate keeping up to date with research evidence to leading or assisting a journal club, reading email subscription alerts, or gaining research-based knowledge from a colleague. This departure from formal sources of research-based knowledge is more aligned with the behaviors and preferences of rehabilitation clinicians who favor informal, quick methods of gaining research evidence, and tend to keep up with research through a variety of informal sources including consultation with trusted peers and email reminders.^15,23,24,47,80–82^ The process by which rehabilitation clinicians rely on colleagues for research-based knowledge is starting to gain importance in the EBP literature as a recognized and beneficial mechanism of EBP.^24,82–84^ Still, this is a promising avenue for future research.

In the third and last step of this study, the effect of language on VAS ratings was trivial, supporting the equivalency of the response option labels in English and French for all items. The distribution of ratings also demonstrates the ordinal consistency of response options and the quasi-interval nature of the scales (i.e., response options are equally spaced). The *keeping up to date* item had the largest variation in ratings between languages and within the same language, however these differences were less than five on 100 and are considered negligeable.^98^ A possible explanation for this variability in ratings may be because of the subjective nature of these behavioral frequency adverbs (potentially dependent on the area of practice) and lack of consensus on how often rehabilitation clinicians should keep up to date with the literature. As knowledge is produced at different rates for different areas of practice, and that this index is meant to be used in various settings, no explicit frequency denominator was attributed to this item. For instance, “regularly” could mean once every two months for a clinician in stroke rehabilitation or once a year for a clinician in palliative care. Our intent with this item is to capture the respondent’s self-rating relative to their understanding of what “regularly”, “occasionally” and “rarely” mean relative to their field of practice and relative to their perception of what is feasible given their clinical reality.

### Limitations

A strength of this study is the multi-phased rigorous qualitative review process which included target end-users and EBP researchers. In developing the response option scaling survey, we aimed to provide adequate guidance to maximize respondents’ comprehension of the task. Pilot testing enabled us to add examples and clarify the instructions. Still, given that 25 individuals did not complete the survey (19%) and that an additional 24 had to be excluded due to apparent miscomprehension of the task (19%), this exercise may have been perceived as difficult and burdensome, a finding also reported by others in the context of a valuation exercise for the EQ-5D.^99^ The task involved an unfamiliar method of placing response option labels on a 0 to 100 scale which required an ability for abstract reasoning. Given the lack of available demographic data, it is impossible to discern who misunderstood the task. Comprehension may have been improved with a quick instructional video.

Before deploying the PIRE-CDMI, there remains an important developmental step which consists of estimating the relative weights of each dimension-level. This work, which the research team has started, will allow for the generation of a more accurate total score that takes into consideration end-users’ perceived relative contribution of dimensions on the overall construct of propensity to integrate research evidence into CDM. We acknowledge that the initial mathematical properties of the prototype PIRE-CDMI established in previous research (Roberge-Dao et al., submitted) may have changed due to item rewriting. While this must be confirmed in future testing, our findings pertaining to the quasi-interval spacing of response option labels gives us reason to believe that the interval properties of the scale still hold. Due to important linguistic differences between different countries, we suggest undergoing a thorough cultural adaptation and reassessment before using the PIRE-CDMI with English and French-speaking individuals outside of Canada.

Finally, while some authors have stated that short scales are a limitation and can compromise the validity and reliability of inferences drawn from a measure,^100,101^ others have found value in the efficiency of short scales.^102,103^ There exists a delicate trade-off between scale comprehensiveness and feasibility. Given the resource-strained healthcare context, the aim was to create a short index capable of rapidly estimating elements of EBP requiring improvement. This measure can be used as an efficient global outcome measure of a clinician’s propensity to integrate research evidence into CDM for research purposes and professional self-reflection which may then be complemented with more comprehensive and lengthier measures. Intervention strategies can then be developed to target the specific areas requiring support.

## Conclusion

The three consecutive phases described in this paper illustrate a rigorous approach to developing a brief multidimensional index of propensity to integrate research evidence into CDM in rehabilitation that is coherent, clear, and relevant to Canadian OTs and PTs. A focus group and cognitive interviews gave rise to important item modifications in English and French to minimize ambiguity, measurement bias and cognitive burden on respondents. Finally, response option labels in English and French were found to be equivalent through a cross-sectional online survey wherein response option labels were compared on 0 to 100 scales in both languages. It is hoped that the use of this practical index will help identify research and practice needs, better support clinicians and improve the quality of rehabilitation care.

## Data Availability

All data produced or analyzed in the present study are available upon reasonable request to the authors

## Acknowledgments

This study was funded by the Canadian Institutes of Health Research (grant number 148544). The funding body did not influence the study results and was not involved in the study in any way.

## Appendices

## Appendix I. Five PIRE-CDMI response sets

| Dimension | Response options | Response anchors |
| --- | --- | --- |
| Use of research evidence | Almost always<br>Sometimes<br>Rarely | None of the time / All of the time |
| Self-efficacy | Confident<br>Somewhat confident<br>Not very confident | No confidence / full confidence |
| Resources | Have the necessary resources<br>Have some of the necessary resources<br>Have few of the necessary resources | None / All imaginable resources |
| Attitudes □ Effort | Little effort<br>Moderate effort<br>A lot of effort | No effort / Full effort |
| Activities related to EBP □ Keeping up to date | Regularly<br>Occasionally<br>Rarely | 0 days/month /<br>30 days/month |

## Appendix II. Step-by-step changes at each step of the item rewriting process in both languages for each item English version of the index (please contact the author for French version of the index)

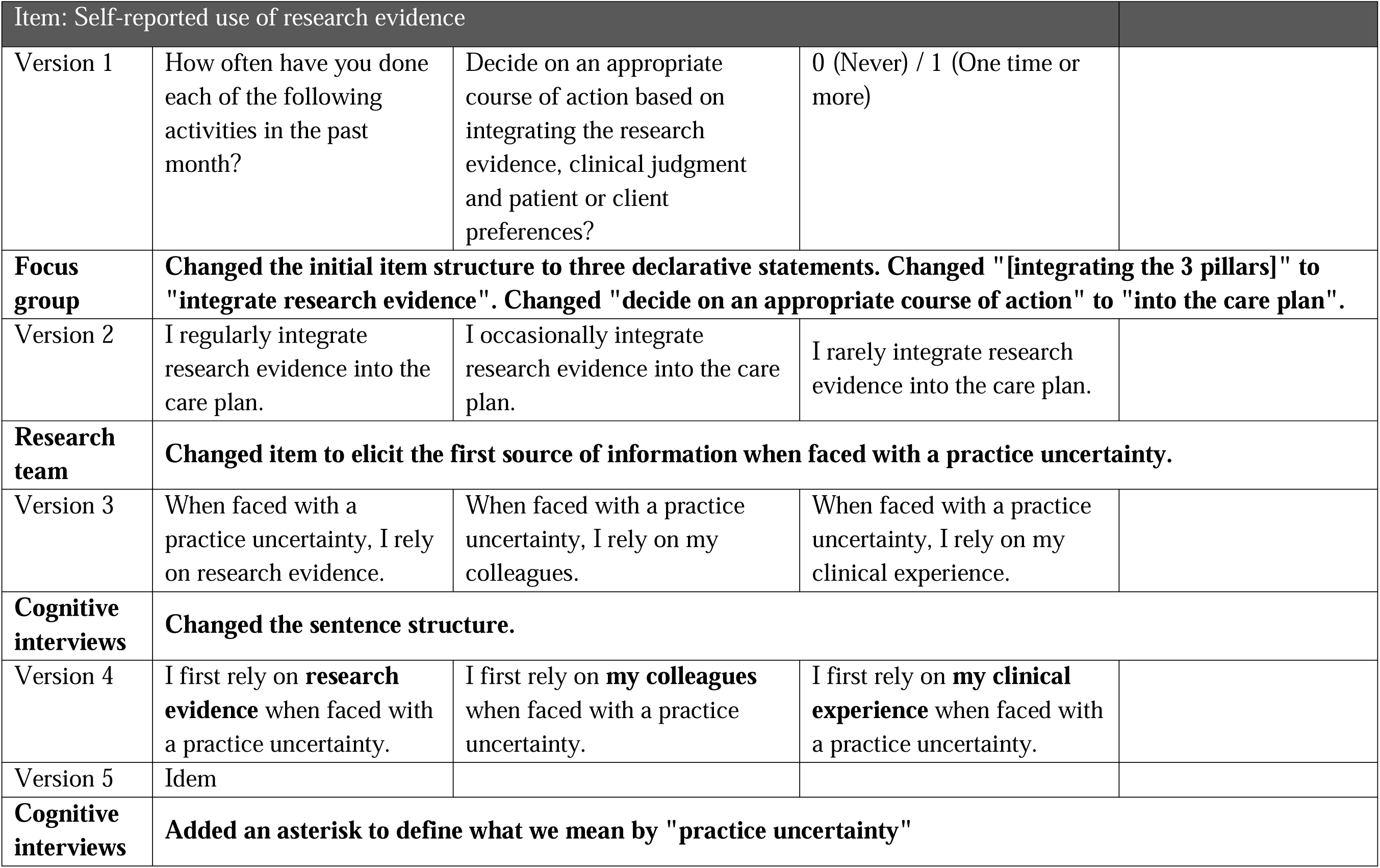
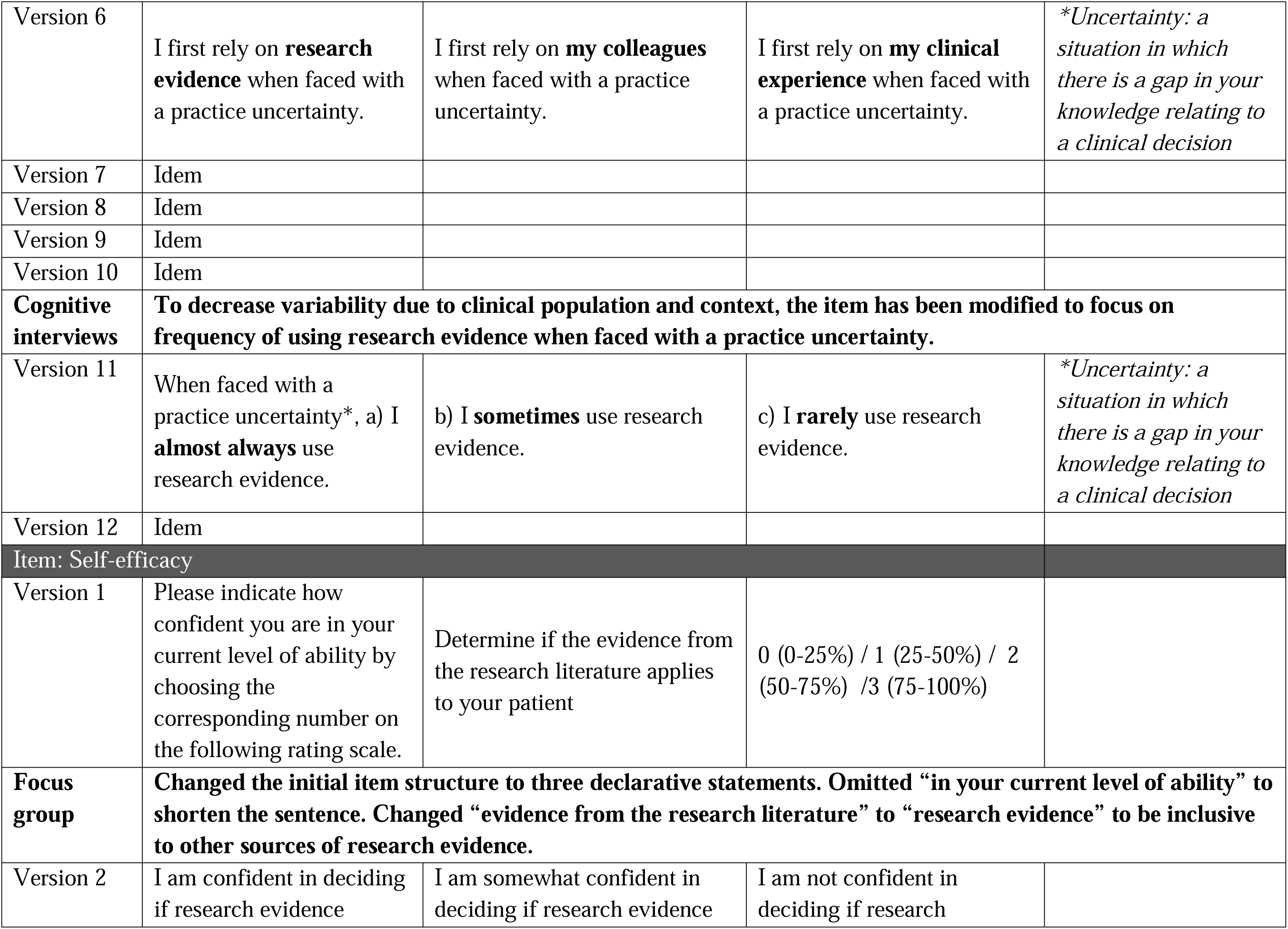
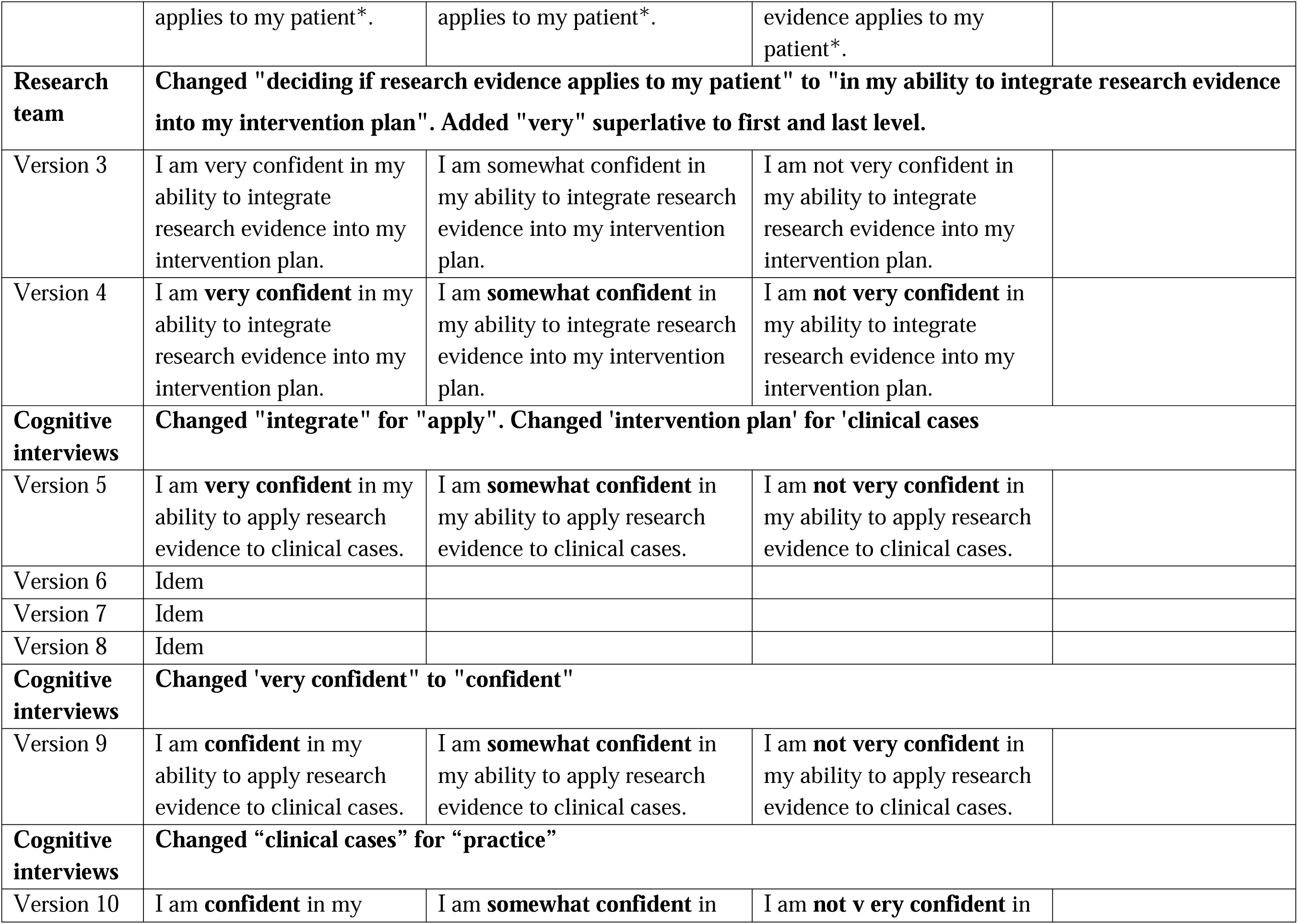
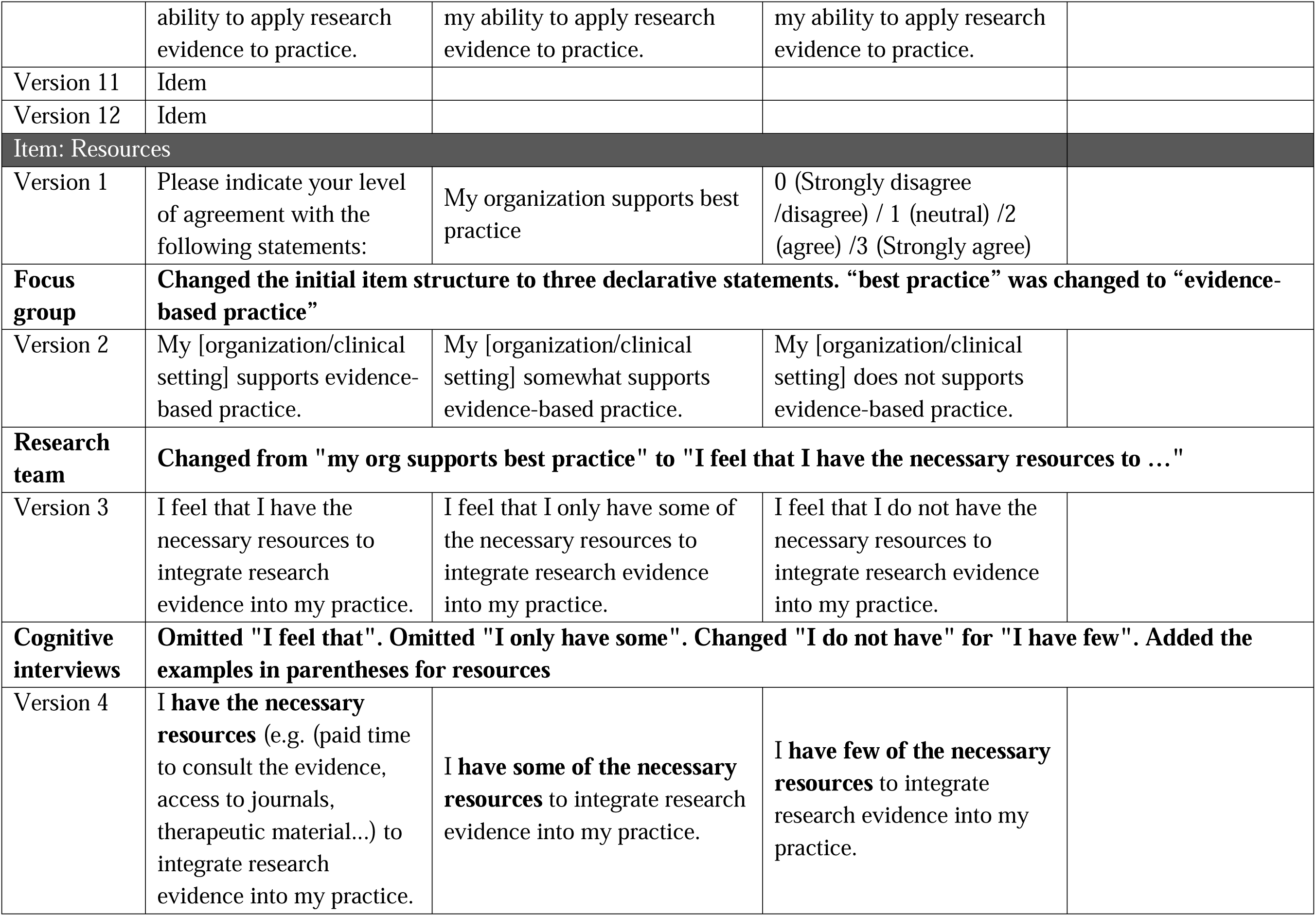
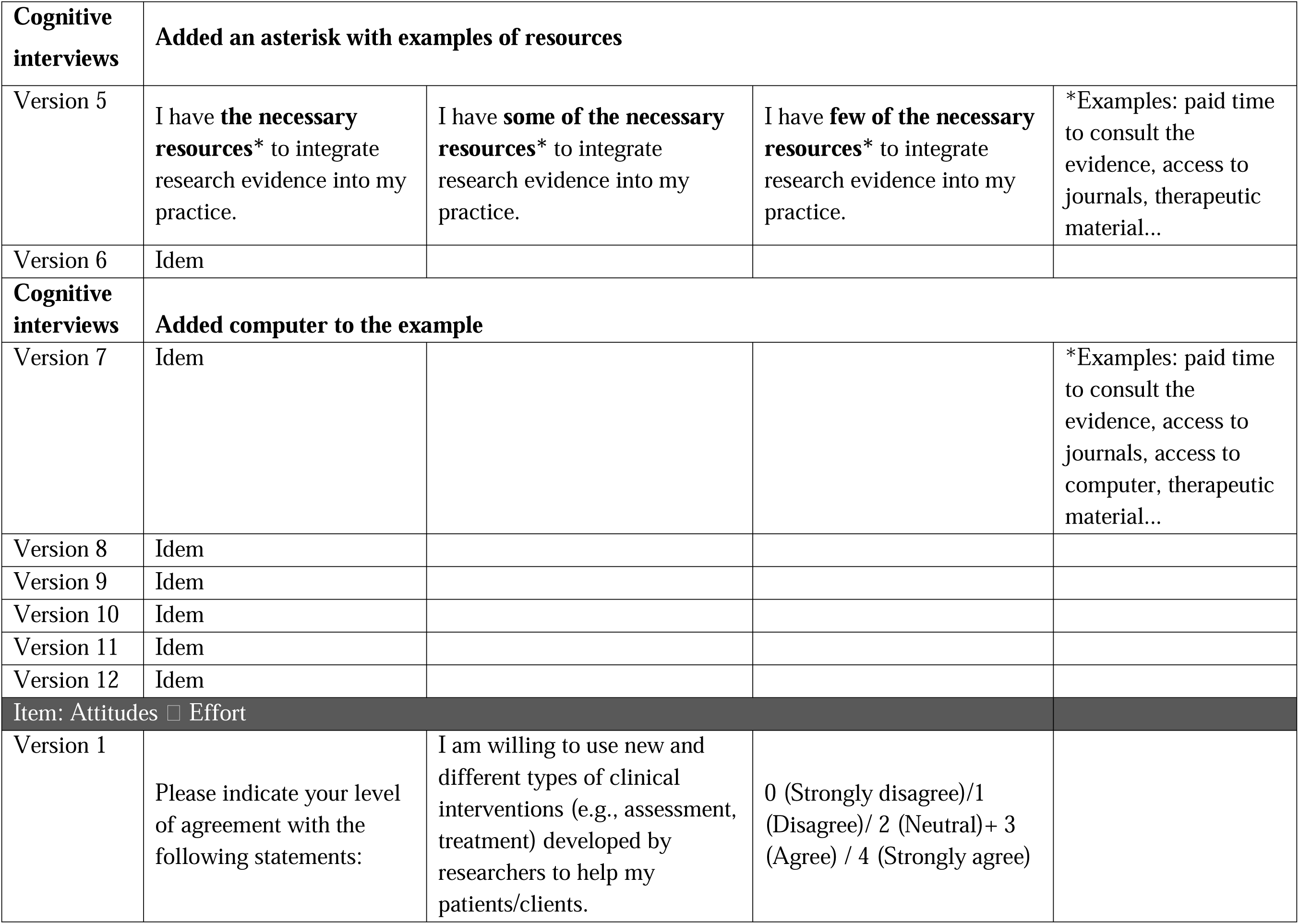
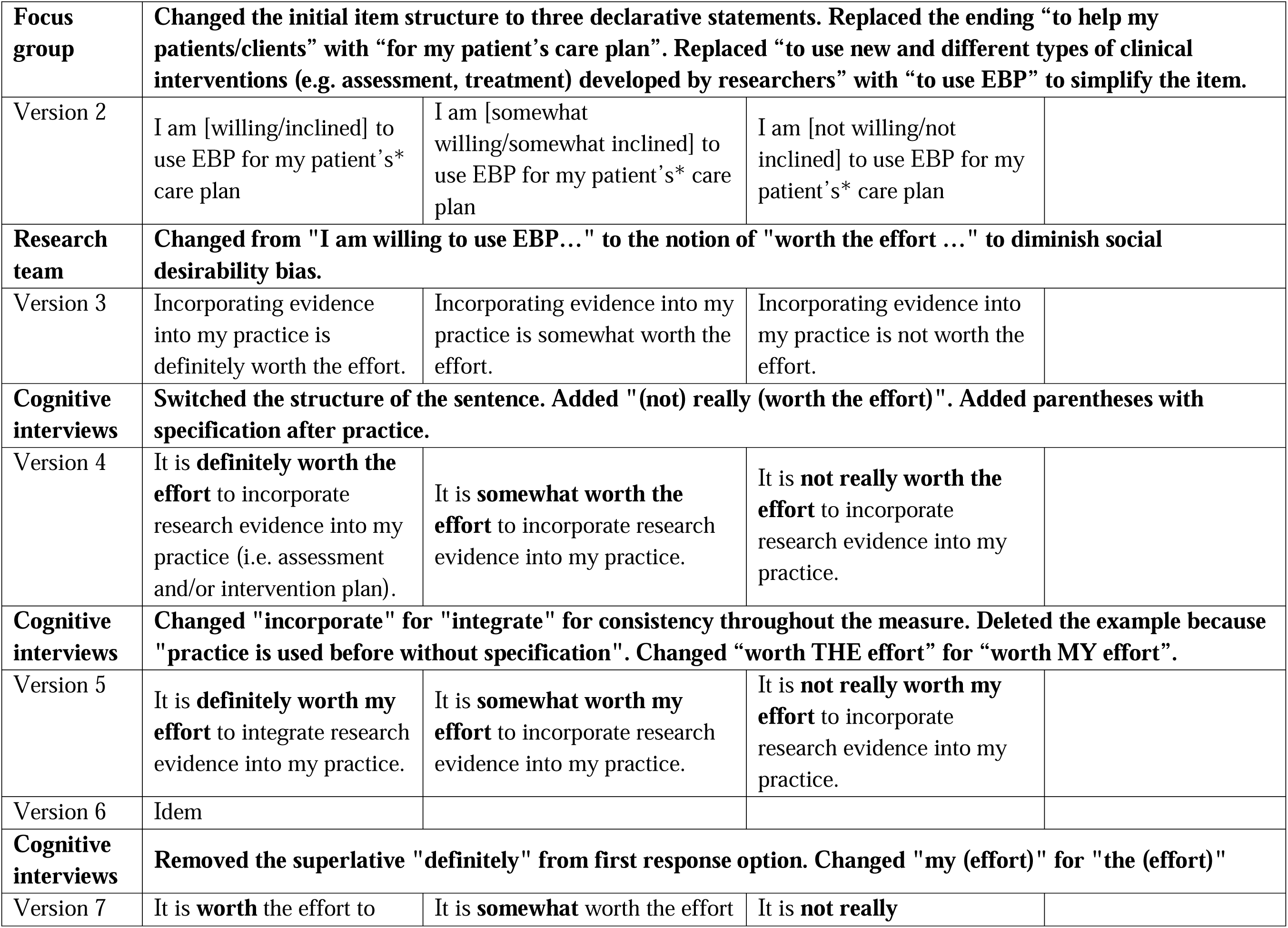
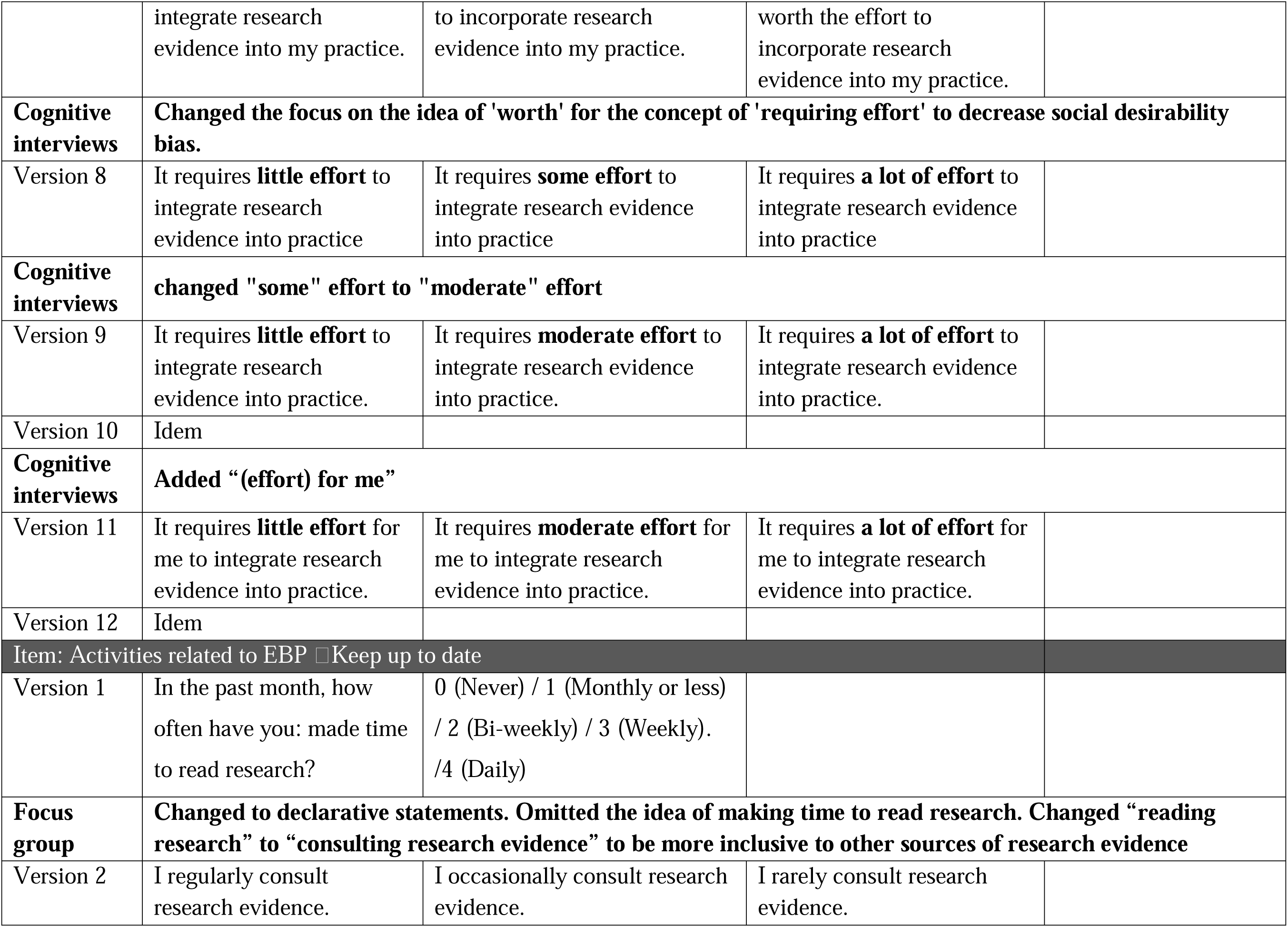
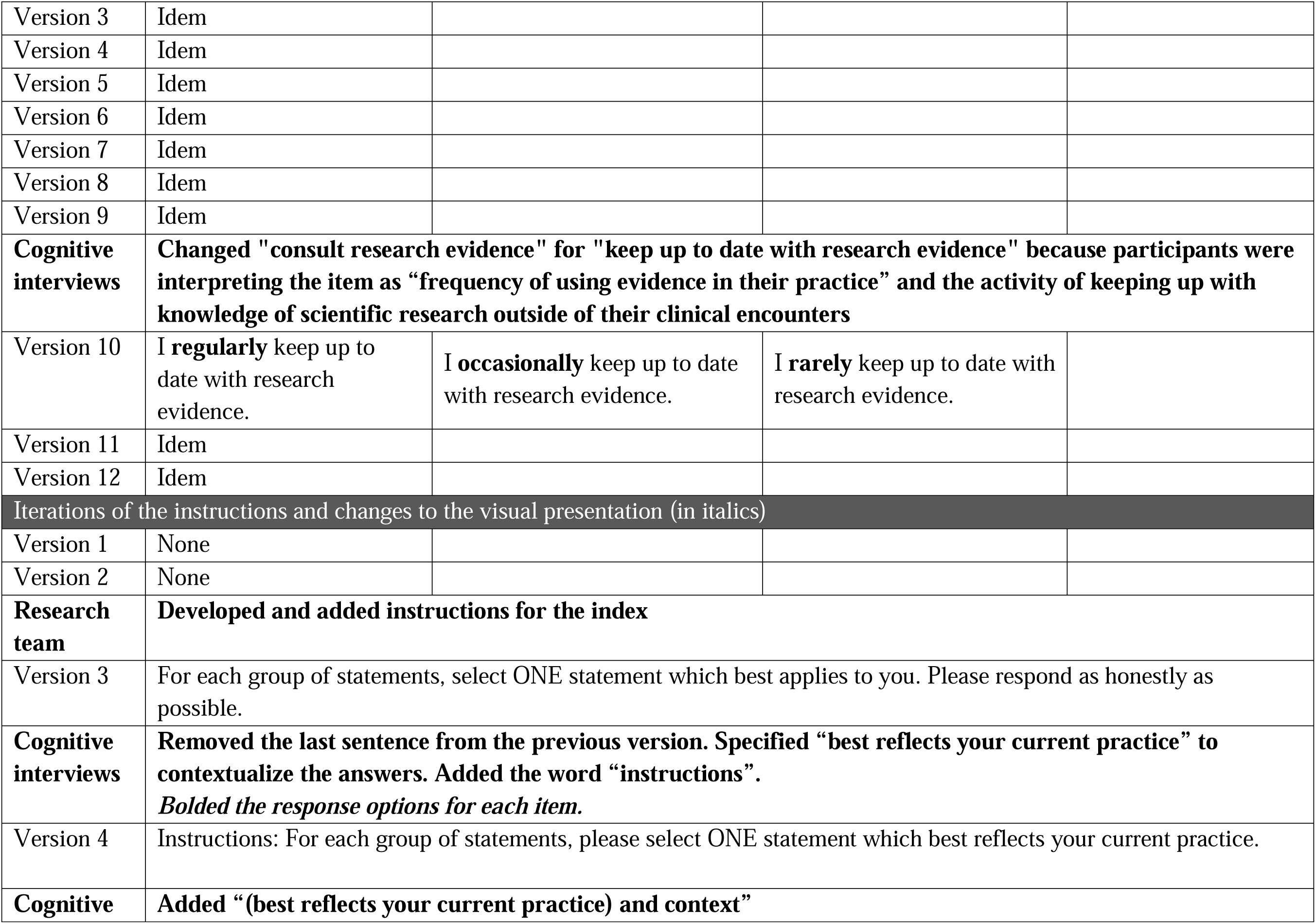
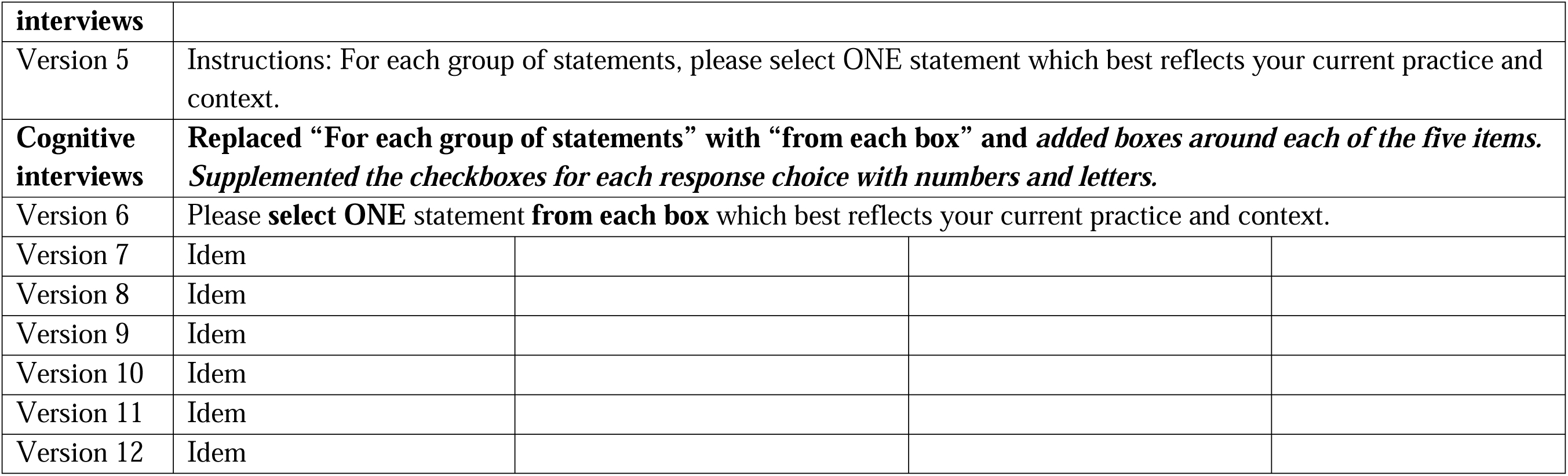

| Item: Self-reported use of research evidence |  |  |  |
| --- | --- | --- | --- |
| Version 1 | How often have you done each of the following activities in the past month? | Decide on an appropriate course of action based on integrating the research evidence, clinical judgment and patient or client preferences? | 0 (Never) / 1 (One time or more) |
| **Focus group** | **Changed the initial item structure to three declarative statements. Changed "[integrating the 3 pillars]" to "integrate research evidence". Changed "decide on an appropriate course of action" to "into the care plan".** |  |  |
| Version 2 | I regularly integrate research evidence into the care plan. | I occasionally integrate research evidence into the care plan. | I rarely integrate research evidence into the care plan. |
| **Research team** | **Changed item to elicit the first source of information when faced with a practice uncertainty.** |  |  |
| Version 3 | When faced with a practice uncertainty, I rely on research evidence. | When faced with a practice uncertainty, I rely on my colleagues. | When faced with a practice uncertainty, I rely on my clinical experience. |
| **Cognitive interviews** | **Changed the sentence structure.** |  |  |
| Version 4 | I first rely on **research evidence** when faced with a practice uncertainty. | I first rely on **my colleagues** when faced with a practice uncertainty. | I first rely on **my clinical experience** when faced with a practice uncertainty. |
| Version 5 | Idem |  |  |
| **Cognitive interviews** | **Added an asterisk to define what we mean by "practice uncertainty"** |  |  |

|  |  |  |  |  |
| --- | --- | --- | --- | --- |
| Version 6 | I first rely on **research evidence** when faced with a practice uncertainty. | I first rely on **my colleagues** when faced with a practice uncertainty. | I first rely on **my clinical experience** when faced with a practice uncertainty. | *\*Uncertainty: a situation in which there is a gap in your knowledge relating to a clinical decision* |
| Version 7 | Idem |  |  |  |
| Version 8 | Idem |  |  |  |
| Version 9 | Idem |  |  |  |
| Version 10 | Idem |  |  |  |
| **Cognitive interviews** | **To decrease variability due to clinical population and context, the item has been modified to focus on frequency of using research evidence when faced with a practice uncertainty.** |  |  |  |
| Version 11 | When faced with a practice uncertainty*, a) I **almost always** use research evidence. | b) I **sometimes** use research evidence. | c) I **rarely** use research evidence. | *\*Uncertainty: a situation in which there is a gap in your knowledge relating to a clinical decision* |
| Version 12 | Idem |  |  |  |
| Item: Self-efficacy |  |  |  |  |
| Version 1 | Please indicate how confident you are in your current level of ability by choosing the corresponding number on the following rating scale. | Determine if the evidence from the research literature applies to your patient | 0 (0-25%) / 1 (25-50%) / 2 (50-75%) / 3 (75-100%) |  |
| **Focus group** | **Changed the initial item structure to three declarative statements. Omitted “in your current level of ability” to shorten the sentence. Changed “evidence from the research literature” to “research evidence” to be inclusive to other sources of research evidence.** |  |  |  |
| Version 2 | I am confident in deciding if research evidence | I am somewhat confident in deciding if research evidence | I am not confident in deciding if research |  |
|  | applies to my patient*. | applies to my patient*. | evidence applies to my patient*. |
| **Research team** | **Changed "deciding if research evidence applies to my patient" to "in my ability to integrate research evidence into my intervention plan". Added "very" superlative to first and last level.** |  |  |
| Version 3 | I am very confident in my ability to integrate research evidence into my intervention plan. | I am somewhat confident in my ability to integrate research evidence into my intervention plan. | I am not very confident in my ability to integrate research evidence into my intervention plan. |
| Version 4 | I am **very confident** in my ability to integrate research evidence into my intervention plan. | I am **somewhat confident** in my ability to integrate research evidence into my intervention plan. | I am **not very confident** in my ability to integrate research evidence into my intervention plan. |
| **Cognitive interviews** | **Changed "integrate" for "apply". Changed 'intervention plan' for 'clinical cases** |  |  |
| Version 5 | I am **very confident** in my ability to apply research evidence to clinical cases. | I am **somewhat confident** in my ability to apply research evidence to clinical cases. | I am **not very confident** in my ability to apply research evidence to clinical cases. |
| Version 6 | Idem |  |  |
| Version 7 | Idem |  |  |
| Version 8 | Idem |  |  |
| **Cognitive interviews** | **Changed 'very confident' to "confident"** |  |  |
| Version 9 | I am **confident** in my ability to apply research evidence to clinical cases. | I am **somewhat confident** in my ability to apply research evidence to clinical cases. | I am **not very confident** in my ability to apply research evidence to clinical cases. |
| **Cognitive interviews** | **Changed “clinical cases” for “practice”** |  |  |
| Version 10 | I am **confident** in my | I am **somewhat confident** in | I am **not v ery confident** in |
|  | ability to apply research evidence to practice. | my ability to apply research evidence to practice. | my ability to apply research evidence to practice. |
| Version 11 | Idem |  |  |
| Version 12 | Idem |  |  |
| Item: Resources |  |  |  |
| Version 1 | Please indicate your level of agreement with the following statements: | My organization supports best practice | 0 (Strongly disagree /disagree) / 1 (neutral) /2 (agree) /3 (Strongly agree) |
| **Focus group** | **Changed the initial item structure to three declarative statements. “best practice” was changed to “evidence-based practice”** |  |  |
| Version 2 | My [organization/clinical setting] supports evidence-based practice. | My [organization/clinical setting] somewhat supports evidence-based practice. | My [organization/clinical setting] does not supports evidence-based practice. |
| **Research team** | **Changed from "my org supports best practice" to "I feel that I have the necessary resources to ..."** |  |  |
| Version 3 | I feel that I have the necessary resources to integrate research evidence into my practice. | I feel that I only have some of the necessary resources to integrate research evidence into my practice. | I feel that I do not have the necessary resources to integrate research evidence into my practice. |
| **Cognitive interviews** | **Omitted "I feel that". Omitted "I only have some". Changed "I do not have" for "I have few". Added the examples in parentheses for resources** |  |  |
| Version 4 | **I have the necessary resources** (e.g. (paid time to consult the evidence, access to journals, therapeutic material...) to integrate research evidence into my practice. | **I have some of the necessary resources** to integrate research evidence into my practice. | **I have few of the necessary resources** to integrate research evidence into my practice. |

|  |  |  |  |  |
| --- | --- | --- | --- | --- |
| **Cognitive interviews** | **Added an asterisk with examples of resources** |  |  |  |
| Version 5 | I have **the necessary resources\*** to integrate research evidence into my practice. | I have **some of the necessary resources\*** to integrate research evidence into my practice. | I have **few of the necessary resources\*** to integrate research evidence into my practice. | *Examples: paid time to consult the evidence, access to journals, therapeutic material... |
| Version 6 | Idem |  |  |  |
| **Cognitive interviews** | **Added computer to the example** |  |  |  |
| Version 7 | Idem |  |  | *Examples: paid time to consult the evidence, access to journals, access to computer, therapeutic material... |
| Version 8 | Idem |  |  |  |
| Version 9 | Idem |  |  |  |
| Version 10 | Idem |  |  |  |
| Version 11 | Idem |  |  |  |
| Version 12 | Idem |  |  |  |
| Item: Attitudes ☐ Effort |  |  |  |  |
| Version 1 | Please indicate your level of agreement with the following statements: | I am willing to use new and different types of clinical interventions (e.g., assessment, treatment) developed by researchers to help my patients/clients. | 0 (Strongly disagree)/1 (Disagree)/ 2 (Neutral)+ 3 (Agree) / 4 (Strongly agree) |  |
| **Focus group** | **Changed the initial item structure to three declarative statements. Replaced the ending “to help my patients/clients” with “for my patient’s care plan”. Replaced “to use new and different types of clinical interventions (e.g. assessment, treatment) developed by researchers” with “to use EBP” to simplify the item.** |  |  |
| Version 2 | I am [willing/inclined] to use EBP for my patient’s* care plan | I am [somewhat willing/somewhat inclined] to use EBP for my patient’s* care plan | I am [not willing/not inclined] to use EBP for my patient’s* care plan |
| **Research team** | **Changed from "I am willing to use EBP..." to the notion of "worth the effort ..." to diminish social desirability bias.** |  |  |
| Version 3 | Incorporating evidence into my practice is definitely worth the effort. | Incorporating evidence into my practice is somewhat worth the effort. | Incorporating evidence into my practice is not worth the effort. |
| **Cognitive interviews** | **Switched the structure of the sentence. Added "(not) really (worth the effort)". Added parentheses with specification after practice.** |  |  |
| Version 4 | It is **definitely worth the effort** to incorporate research evidence into my practice (i.e. assessment and/or intervention plan). | It is **somewhat worth the effort** to incorporate research evidence into my practice. | It is **not really worth the effort** to incorporate research evidence into my practice. |
| **Cognitive interviews** | **Changed "incorporate" for "integrate" for consistency throughout the measure. Deleted the example because "practice is used before without specification". Changed “worth THE effort” for “worth MY effort”.** |  |  |
| Version 5 | It is **definitely worth my effort** to integrate research evidence into my practice. | It is **somewhat worth my effort** to incorporate research evidence into my practice. | It is **not really worth my effort** to incorporate research evidence into my practice. |
| Version 6 | Idem |  |  |
| **Cognitive interviews** | **Removed the superlative "definitely" from first response option. Changed "my (effort)" for "the (effort)"** |  |  |
| Version 7 | It is **worth** the effort to | It is **somewhat** worth the effort | It is **not really** |
|  | integrate research evidence into my practice. | to incorporate research evidence into my practice. | worth the effort to incorporate research evidence into my practice. |
| **Cognitive interviews** | **Changed the focus on the idea of 'worth' for the concept of 'requiring effort' to decrease social desirability bias.** |  |  |
| Version 8 | It requires **little effort** to integrate research evidence into practice | It requires **some effort** to integrate research evidence into practice | It requires **a lot of effort** to integrate research evidence into practice |
| **Cognitive interviews** | **changed "some" effort to "moderate" effort** |  |  |
| Version 9 | It requires **little effort** to integrate research evidence into practice. | It requires **moderate effort** to integrate research evidence into practice. | It requires **a lot of effort** to integrate research evidence into practice. |
| Version 10 | Idem |  |  |
| **Cognitive interviews** | **Added “(effort) for me”** |  |  |
| Version 11 | It requires **little effort** for me to integrate research evidence into practice. | It requires **moderate effort** for me to integrate research evidence into practice. | It requires **a lot of effort** for me to integrate research evidence into practice. |
| Version 12 | Idem |  |  |
| **Item: Activities related to EBP ☐ Keep up to date** |  |  |  |
| Version 1 | In the past month, how often have you: made time to read research? | 0 (Never) / 1 (Monthly or less) / 2 (Bi-weekly) / 3 (Weekly). /4 (Daily) |  |
| **Focus group** | **Changed to declarative statements. Omitted the idea of making time to read research. Changed “reading research” to “consulting research evidence” to be more inclusive to other sources of research evidence** |  |  |
| Version 2 | I regularly consult research evidence. | I occasionally consult research evidence. | I rarely consult research evidence. |
| Version 3 | Idem |  |  |
| Version 4 | Idem |  |  |
| Version 5 | Idem |  |  |
| Version 6 | Idem |  |  |
| Version 7 | Idem |  |  |
| Version 8 | Idem |  |  |
| Version 9 | Idem |  |  |
| **Cognitive interviews** | **Changed "consult research evidence" for "keep up to date with research evidence" because participants were interpreting the item as “frequency of using evidence in their practice” and the activity of keeping up with knowledge of scientific research outside of their clinical encounters** |  |  |
| Version 10 | I **regularly** keep up to date with research evidence. | I **occasionally** keep up to date with research evidence. | I **rarely** keep up to date with research evidence. |
| Version 11 | Idem |  |  |
| Version 12 | Idem |  |  |
| Iterations of the instructions and changes to the visual presentation (in italics) |  |  |  |
| Version 1 | None |  |  |
| Version 2 | None |  |  |
| **Research team** | **Developed and added instructions for the index** |  |  |
| Version 3 | For each group of statements, select ONE statement which best applies to you. Please respond as honestly as possible. |  |  |
| **Cognitive interviews** | **Removed the last sentence from the previous version. Specified “best reflects your current practice” to contextualize the answers. Added the word “instructions”.**<br>***Bolded the response options for each item.*** |  |  |
| Version 4 | Instructions: For each group of statements, please select ONE statement which best reflects your current practice. |  |  |
| **Cognitive** | **Added “(best reflects your current practice) and context”** |  |  |
| **interviews** |  |  |  |
| Version 5 | Instructions: For each group of statements, please select ONE statement which best reflects your current practice and context. |  |  |
| **Cognitive interviews** | **Replaced “For each group of statements” with “from each box” and *added boxes around each of the five items.* Supplemented the checkboxes for each response choice with numbers and letters.** |  |  |
| Version 6 | Please **select ONE** statement **from each box** which best reflects your current practice and context. |  |  |
| Version 7 | Idem |  |  |
| Version 8 | Idem |  |  |
| Version 9 | Idem |  |  |
| Version 10 | Idem |  |  |
| Version 11 | Idem |  |  |
| Version 12 | Idem |  |  |

## Appendix III. Overview of the item evolution process during cognitive interviews

| Participant # | Language | Profession | Index version | Use of RE | Self-efficacy | Resources | Attitudes | Activities related to EBP | Instructions | Visual |
| --- | --- | --- | --- | --- | --- | --- | --- | --- | --- | --- |
| P1 | EN | PT | 3 | Major | Minor | Major | Minor |  | Minor | Minor |
| P2 | EN | PT | 3 | Minor |  |  | Minor |  | Minor |  |
| P3 | EN | OT | 3 | Major |  | Minor | Minor |  | Minor |  |
| P4 | FR | OT | 3 | Minor | Major | Minor | Minor |  | Minor |  |
| *New version 4* |  |  |  | **M** |  | **M** | **M** |  | **M** | **M** |
| P5 | EN | OT | 4 |  | Minor | Minor |  |  |  |  |
| *New version 5* |  |  |  |  | **M** | **M** | **M** |  | **M** |  |
| P6 | FR | OT | 5 | Minor |  | Minor |  |  |  |  |
| P7 | EN | OT | 5 | Minor |  |  |  |  | Minor | Minor |
| P8 | EN | PT | 5 | Minor | Minor |  | Major |  | Minor |  |
| *New version 6* |  |  |  | **M** |  |  |  |  | **M** | **M** |
| P9 | FR | PT | 6 |  |  |  | Minor | Minor |  |  |
| P10 | FR | OT | 6 |  |  | Minor | Minor |  |  |  |
| *New version 7* |  |  |  |  |  | **M** | **M** |  |  |  |
| P11 | FR | PT | 7 |  | Minor |  | Major |  |  |  |
| P12 | FR/EN | OT | 7 |  | Minor | Minor | Minor |  |  |  |
| P13 | FR | PT | 7 | Minor |  |  | Major |  |  |  |
| *New version 8* |  |  |  |  |  |  | **M** |  |  |  |
| P14 | EN | PT | 8 | Minor |  |  | Minor |  |  |  |
| P15 | EN | OT | 8 |  | Minor |  | Minor |  |  |  |
| *New version 9* |  |  |  |  | **M** |  | **M** |  |  |  |
| P16 | EN | OT | 9 |  | Minor |  |  | Minor |  |  |
| P17 | FR | PT | 9 |  |  |  | Minor | Minor |  |  |
| *New version 10* |  |  |  |  | **M** |  |  | **M** |  |  |
| P18 | FR | OT | 10 |  |  |  |  |  |  |  |
| P19 | FR | OT | 10 | Major |  |  | Minor | Minor |  |  |
| P20 | FR | OT | 10 | Minor | Minor | Minor |  |  |  |  |
| P21 | EN | OT | 10 | Minor |  |  |  |  |  |  |
| *New version 11* |  |  |  | **M** |  |  | **M** |  |  |  |
| P22 | FR | PT | 11 |  |  |  |  |  |  |  |
| P23 | EN | OT | 11 |  |  |  |  |  |  |  |
| P24 | EN | PT | 11 |  |  |  |  | Minor |  |  |
| *Version 12 accepted* |  |  |  |  |  |  |  |  |  |  |
| Total iterations during cognitive interviews |  |  |  | 3 | 3 | 3 | 6 | 1 |  |  |

EN: English; FR: French; PT: physical therapist; OT: occupational therapist; RE: research evidence; Major: Major issue identified by participants; Minor: minor issue identified by participants; M: Modification implemented by the research team into next version of the index

## Appendix IV. Final index in English (please contact the author for the Final index in French)

**Propensity to Integrate Research Evidence into Clinical Decision-Making Index (PIRE-CDMI)**

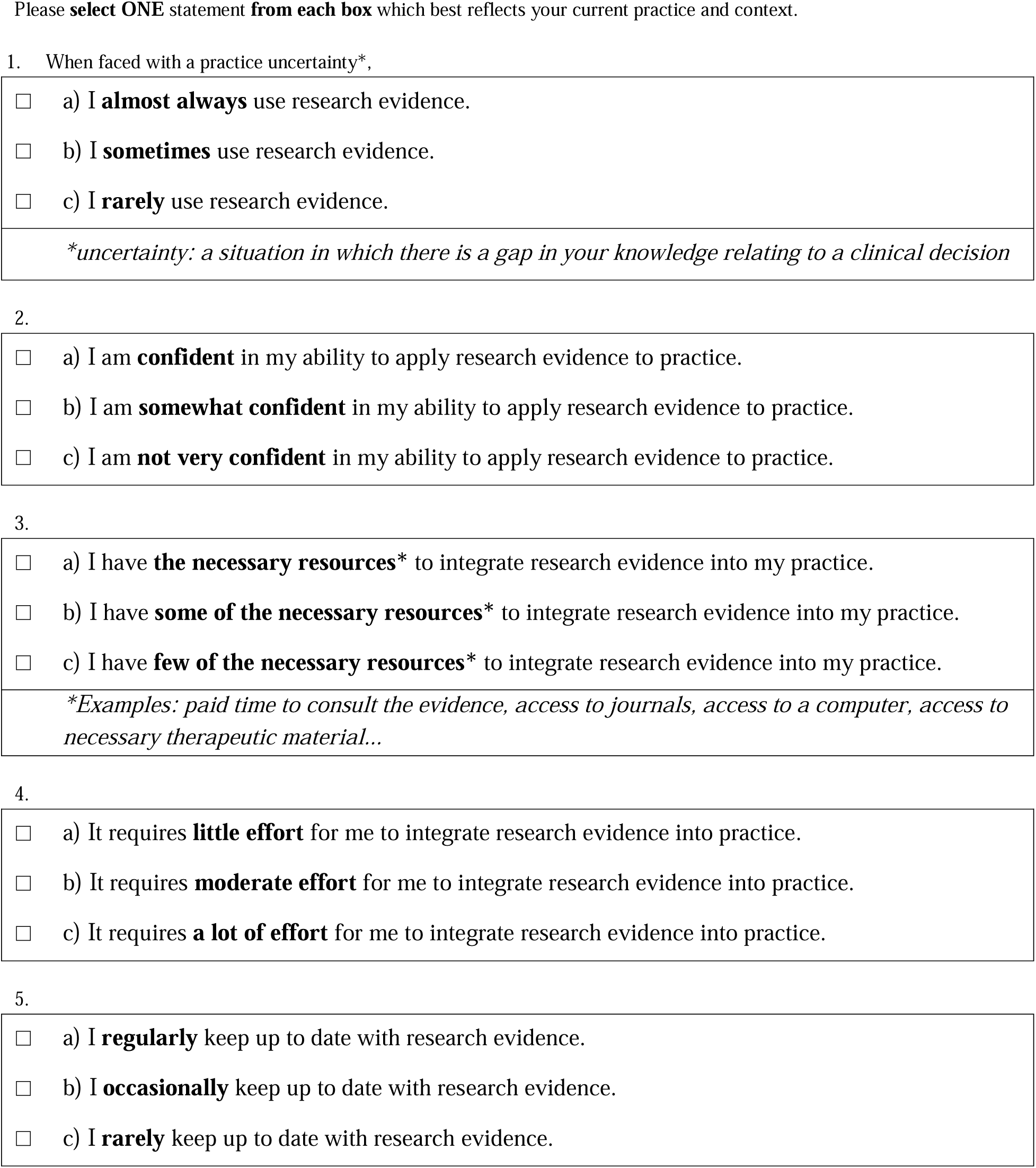

